# Disrupted balance between pro-inflammatory lipid mediators and anti-inflammatory specialized pro-resolving mediators is linked to hyperinflammation in patients with alcoholic hepatitis

**DOI:** 10.1101/2023.12.15.23300034

**Authors:** Wei Li, Ying Xia, Jing Yang, Arun J. Sanyal, Vijay H. Shah, Naga P. Chalasani, Qigui Yu

## Abstract

**Background:** Chronic excessive alcohol consumption leads to a spectrum of alcohol-associated liver diseases (ALD), including alcoholic hepatitis (AH). AH is characterized by intense systemic and liver inflammation, posing significant risks of health complications and mortality. While inflammation is a crucial defense mechanism against injury and infection, its timely resolution is essential to prevent tissue damage and restore tissue homeostasis. The resolution of inflammation is an actively regulated process, primarily governed by specialized pro-resolving mediators (SPMs), lipid metabolites derived from ω-6 and ω-3 poly-unsaturated fatty acids (PUFAs). We investigated the balance between pro-inflammatory lipid mediators (PLMs) and SPMs in the ω-6 and ω-3 PUFA metabolic pathways and examined the impact of alcohol abstinence on rectifying the dysregulated biosynthesis of PLMs and SPMs in AH patients.

**Methods:** LC-MS/MS and ELISA were used to quantify levels of bioactive lipid mediators (LMs) and their precursors in the plasma samples from 58 AH patients, 29 heavy drinkers without overt liver diseases (HDCs), and 35 healthy controls (HCs). Subsequently, we assessed correlations of altered LMs with clinical parameters and various markers of inflammatory cascade andmicrobial translocation. Furthermore, we conducted a longitudinal study to track changes in levels of LMs over 6- and 12-month follow-ups in AH patients who underwent alcohol abstinence.

**Results:** AH patients exhibited significantly higher plasma levels of ω-6 PLMs (PGD_2_ and LTB_4_) and SPM RvE1 compared to HDCs and/or HCs. Conversely, key SPMs such as LXA4, RvD1, and several precursors in the ω-3 pathway were significantly downregulated in AH patients. Some of these altered LMs were found to correlate with AH disease severity, clinical parameters, and various inflammatory cytokines. In particular, the LTB4/LXA4 ratio was substantially elevated in AH patients relative to HDCs and HCs. This altered ratio displayed a positive correlation with the MELD score, suggesting its potential utility as an indicator of disease severity in AH patients. Importantly, the majority of dysregulated LMs, particularly PLMs, were normalized following alcohol abstinence.

**Conclusion:** Our study reveals significant dysregulation in the levels of PLM metabolites and anti-inflammatory SPMs in both ω-6 and ω-3 PUFA pathways in AH patients. This disrupted biosynthesis, characterized by an overabundance of PLMs and a deficiency in SPMs, is linked to the heightened inflammation observed in AH patients. Importantly, our findings suggest an important role of alcohol abstinence in restoring the balance of these LMs and the potential therapeutic benefits of SPM supplements in alleviating the inflammatory cascade in AH patients.

## Introduction

Long-term heavy alcohol consumption leads to a spectrum of alcohol-associated liver diseases (ALD), including steatosis, alcoholic hepatitis (AH), and liver fibrosis/cirrhosis. AH, a critical and progressive acute-on-chronic liver disorder, is characterized by heightened hepatic and systemic inflammation, and is associated with significant morbidity and mortality. Circulating levels of various inflammatory mediators, such as proinflammatory cytokines (TNF-α, IL-6, and IL-8), are profoundly elevated in AH patients and directly correlate with disease severity and mortality rates^1–4^. These inflammatory mediators primarily originate from persistently hyperactivated immune cells present in both the peripheral blood and the liver of AH patients^4–6^. Alcohol disrupts gap junction integrity of gut mucosal epithelial cells, leading to increased permeability of the gastrointestinal (GI) tract and translocation of microbial components such as lipopolysaccharides (LPS) from the GI tract into the bloodstream and liver^7–11^. Alcohol-induced microbial translocation (MT) has been recognized as a major driver of hepatic and systemic inflammation and immune activation in AH patients^8,9,12,13^. Additionally, alcoholic metabolites such as acetate, reactive oxygen species (ROS), and acetaldehyde can directly provoke inflammation^14^. These inflammatory responses, combined with hyperimmune activation, play a central role in the development of liver fibrosis/cirrhosis, multiple organ failure, and death in AH patients.

While inflammation is a vital defense mechanism against injury and infection, its timely resolution is crucial to prevent tissue damage and restore tissue homeostasis. The inflammatory response comprises two distinct phases: initiation and resolution. In the initiation phase, leukocytes are recruited to injured tissue in response to inflammatory cues, further amplifying the inflammatory response by producing a plethora of proinflammatory mediators. Resolution of inflammation is an actively regulated process, largely governed by a group of bioactive lipids called specialized pro-resolving mediators (SPMs), including lipoxins (LXs), resolvins (Rvs), protectins, and maresins (MaRs). These SPMs are synthesized from long-chain omega-3 (ω-3) and ω-6 polyunsaturated fatty acids (PUFAs) that are sourced from dietary elements such as meat, eggs, and fish oils^15–18^, or derived from ingested short-chain ω-3 (α-linolenic acid, ALA) and ω-6 (linolenic acid, LA) PUFAs from vegetables, plant oils, and seeds^19^. Notably, the long-chain ω-6 PUFA, arachidonic acid (AA), serves as a precursor to several potent pro-inflammatory lipid mediators (PLMs), including prostaglandins (PGs), leukotrienes (LTs), and thromboxanes (Txs). AA is converted by cyclooxygenase (COX)-1 and -COX-2 to PGs (PGD2 and PGE2) and Txs by 5-lipoxygenase (5-LOX) to LTs (e.g. LTB4), leading to initiation of the inflammatory response^15–18^. AA can also be converted by 5-, 12-, and 15-LOX to anti-inflammatory LXs (LXA4 and LXB4). On the other hand, long-chain ω-3 PUFAs such as eicosatetraenoic acid (EPA) and docosahexaenoic acid (DHA) primarily give rise to anti-inflammatory SPMs. EPA ω-3 PUFAs is converted to the E-series Rvs (RvE1 and RvE2) by 15-LOX and 5-LOX^15–18^, while DHA ω-3 PUFA is converted by 15-LOX and 5-LOX to D-series Rvs (RvD1-6) and protectins (PD1, PDX), and by 12-LOX/15-LOX type 1 to MaRs (MaR1 and MaR2)^15–18^. Importantly, the gene expression, activity, and intracellular location of these enzymes can be regulated by a variety of factors such as inflammatory mediators^20–23^, leading to different biosynthetic outcomes of SPMs versus PLMs^17^.

SPMs have been detected in both the circulation and tissues of humans in health and disease^24–32^. SPMs actively trigger cardinal signals for inflammation resolution through (1) counter-regulation of proinflammatory mediators, (2) promotion of polymorphonuclear cell/neutrophil (PMN) clearance, (3) promotion of phagocytosis to eliminate apoptotic cells and cellular debris, (4) production enhancement of anti-inflammatory cytokines such as IL-10, (5) mitigation of oxidative stress (OS), and (6) modulation of immune cell activation^15,16,18,20,33–45^. However, it remains largely unexplored whether the biosynthetic balance between PLMs and SPMs is disrupted in AH patients, potentially contributing to hyperinflammation, and whether alcohol abstinence restores this balance, thereby reversing hyperinflammation, as AH is usually reversible if individuals successfully abstain from alcohol or undergo medical interventions^46^. Alcohol abuse causes AH only in a subset of long-term heavy drinkers. It is not known whether an imbalance between PLM and SPM production contributes to the immunological differences underlining the apparent differential susceptibility to AH. In this study, we assessed the profiles of circulating PLMs and SPMs in AH patients, comparing them with matched heavy drinkers without overt liver diseases (HDCs) and healthy controls (HCs)^4^. Additionally, we monitored changes over 6- and 12-month follow-up intervals of alcohol abstinence. Furthermore, we correlated the altered LMs with patients’ clinical parameters, disease severity, levels of inflammatory mediators, as well as markers of MT such as LPS, soluble CD14 (sCD14), and soluble CD163 (sCD163), along with the systemic inflammation marker (C-reactive protein, CRP). To gain deeper insights, we performed data mining to analyze the expression of genes encoding critical enzymes involved in the biosynthesis of LMs in the peripheral blood and liver tissue of AH patients and HCs.

## Materials and Methods

### Study subjects and blood samples

The study subjects, including 58 AH patients and 29 HDCs at baseline as well as 13 and 9 abstinent AH patients at 6- and 12-month follow-ups, respectively, were part of a cohort of well-characterized AH and age-, gender-, and race-matched individuals with comparable history of heavy drinking but no overt clinical evidence of liver disease enrolled into the multicenter prospective observatory Translational Research and Evolving Alcoholic Hepatitis Treatment 001 study (TREAT 001, NCT02172898). Demographic and clinical characterizations as well as drinking patterns of the study subjects are shown in Table 1 and Supplementary Table 1.

**Table 1.**
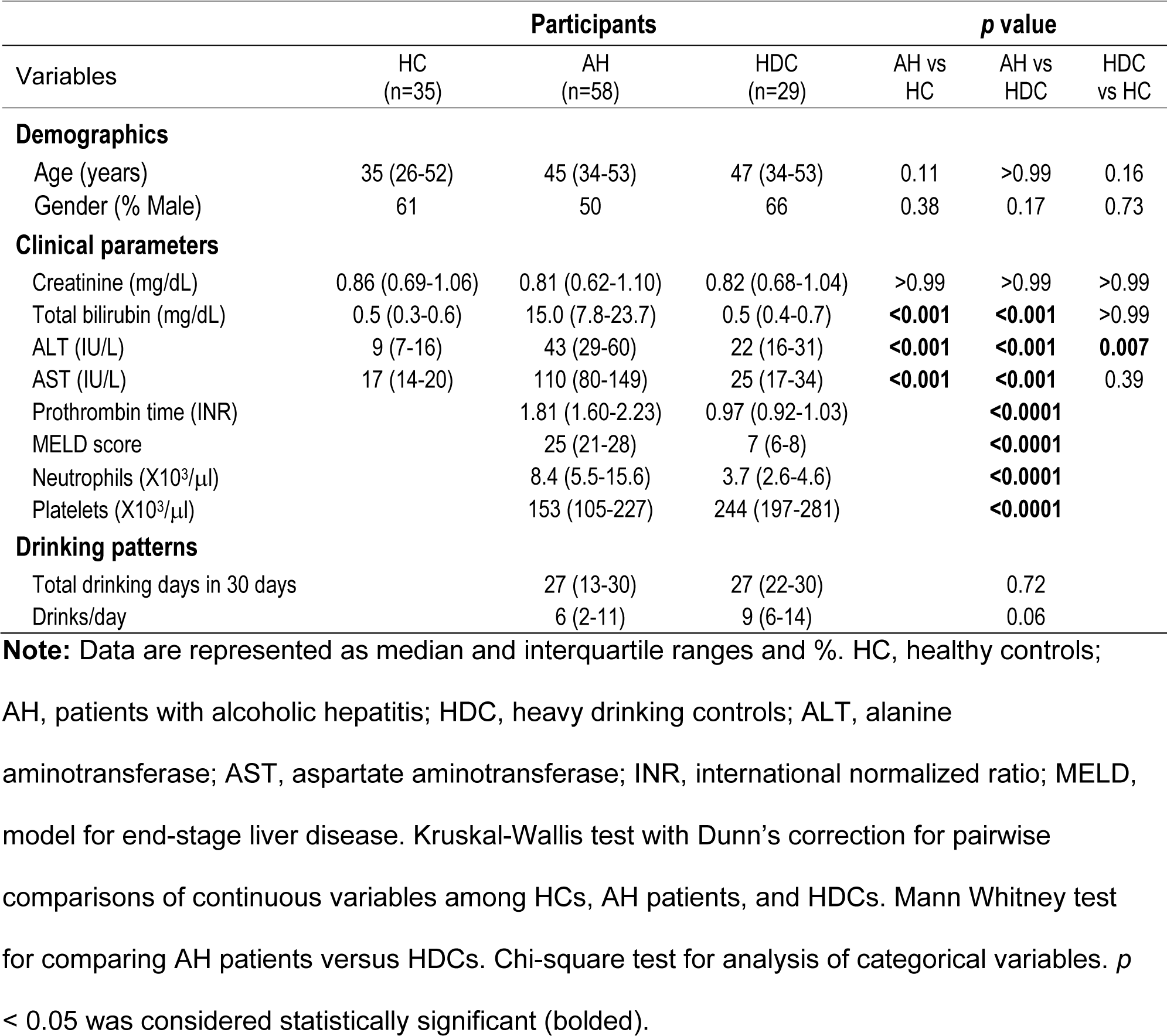
Characteristics of the study cohort.

Detailed definitions of AH and HDC and the inclusion and exclusion criteria were previously described^47^. Briefly, AH was defined as the onset of aspartate aminotransferase (AST) >50 IU/L and elevated total bilirubin (initially >2 mg/dL, later amended to >3 mg/dL) in chronic alcoholics who were drinking heavily within the 6 weeks before recruitment. AH diagnosis in subjects with uncertainty drinking history or atypical clinical features was confirmed by performing a liver biopsy. HDC were age- and gender-matched participants with a similar history of heavy alcohol consumption as the AH patients but had no overt clinical liver disease (AST <50 U/L, alanine aminotransferase [ALT] <50 U/L, total bilirubin within normal range). This study was approved by the Institutional Review Boards (IRB) at Indiana University School of Medicine, Mayo Clinic, and Virginia Commonwealth University. All participants provided a written informed consent form before blood was drawn. Peripheral blood was collected in heparin-coated tubes (BD Biosciences, Franklin Lakes, NJ). Plasma was prepared within 2 hours of blood collection and stored at −80°C until use. Plasma samples from 35 age- and sex -matched healthy volunteers were included as HCs.

### Liquid chromatography with tandem mass spectrometry (LC-MS/MS)

Plasma samples from 10 AH patients, 10 HDCs, and 10 HCs were subjected to LC-MS/MS analysis for the quantitative assessment of PLMs and SPMs. This analytic work was conducted at the Center for Salivary Diagnostics, the Forsyth Institute, Harvard School of Dental Medicine (Cambridge, MA). Briefly, each plasma sample (1 mL) was mixed with internal labeled standards such as d8-5S-HETE, d4-LTB4, d5-LXA4, d5-RvD2, and d4-PGE2 in ice-cold methanol to facilitate the calculation of quantification and sample recovery. The mixtures were subjected to solid phase extraction using C18 cartridges. Extracts were dried using the automated evaporate system (TurboVap, Charlotte, NC), and immediately used for LC-MS/MS automated injections. The LC-MS-MS system, a Shimadzu LC-20AD HPLC and a Shimadzu SIL-20AC autoinjector (Shimadzu, Kyoto, Japan), paired with a QTrap 6500 (ABSciex, Framingham, MA), were employed to process all plasma samples. PLMs, SPM intermediates, and SPMs were identified in accordance with published criteria^17,48,49^, including matching retention time (RT) and at least six characteristic and diagnostic ions^48^. Quantitation was carried out using linear regression compared with standard curves from the synthetic and authentic solvents. LC-MS/MS data analysis was performed on the Sciex software platform, Analyst version 1.6 (Sciex, Framingham, MA)^50^.

### Enzyme-linked immunosorbent assay (ELISA) and multiplex immunoassay

Plasma levels of PGD2, PGE2, LTB4, LXA4, RvE1, RvD2, and MaR1 were quantified using the Prostaglandin D2 ELISA Kit (Cayman Chemical, Ann Arbor, MI), Prostaglandin E2 ELISA Kit (Cayman Chemical, Ann Arbor, MI), LTB4 Parameter Assay Kit (R&D Systems, Minneapolis, MN), Lipoxin A4 ELISA Kit (Neogen, Lexington, KY), Human Resolvin E1 ELISA Kit (MyBioSource, San Diego, CA), Resolvin D2 ELISA Kit (Cayman Chemical, Ann Arbor, MI), and Maresin 1 ELISA Kit (Cayman Chemical, Ann Arbor, MI), respectively. Plasma levels of the systematic inflammation marker CRP and the bacterial translocation markers, including LPS binding protein (LBP) and soluble CD14 (sCD14), were measured using the Human C-Reactive Protein/CRP DuoSet ELISA Kit, Human LBP DuoSet ELISA Kit, and Human CD14 Quantikine ELISA Kit, respectively. These ELISA kits were purchased from R&D Systems (Minneapolis, MN).

Plasma levels of 45 inflammatory mediators, including 26 cytokines, 8 chemokines, and 11 growth factors were simultaneously quantified using the Cytokine/Chemokine/Growth Factor 45-Plex Human ProcartaPlex Panel 1 (EPX450-12171-901, Invitrogen, Waltham, MA), as previously described^51^. The concentrations of these cytokines/chemokines/growth factors were calculated using the Bio-Plex Manager v6.1 software (Bio-Rad, Hercules, CA). For statistical analyses, values below the detection limit of the assay were replaced with the minimal detectable concentrations for each analyte as provided by the manufacturer.

### Data mining of public RNA-seq datasets of liver tissue, monocytes, and neutrophils from AH patients and HCs

Data mining was performed to examine differential expression of genes involved in production of LMs in liver tissue from AH patients and HCs. RNA expression levels were pooled from 3 liver tissue RNA-seq databases GSE143318 (AH, n=5; HC, n=5), GSE142530 (AH, n=10; HC, n=12), and GSE155907 (AH, n=5; HC, n=4). RNA expression of genes involved in LM production in peripheral blood CD14^+^ monocytes from patients with severe AH (n=4) and HCs (n=6) was obtained from the RNA-seq dataset GSE135285. RNA expression levels were also extracted from the neutrophil RNA-seq database GSE1710809 to assess differential expression of genes involved in production of LMs in peripheral blood neutrophils in AH patients (n=3) and HCs (n=3). Data are presented as transcripts per kilobase million (TPM) normalized expression values.

### Statistical analysis

Chi-square test was used for comparison between groups for categorical variables. Mann-Whitney test and Kruskal-Wallis test with Dunn’s corrections were used to calculate differences in continuous variables between 2 groups and among 3 groups in cross-sectional analysis, respectively. The linear relationship between LMs and clinical parameters or inflammatory mediators was analyzed using the Spearman correlation test. Wilcoxon matched-pairs signed rank test was used to calculate the differences in longitudinal analysis. *p* <0.05 was considered statistically significant.

## Results

### Characteristics of the study cohort

This study cohort included 58 AH patients, 29 HDCs, and 35 HCs at baseline, as well as 13 and 9 alcohol abstinent AH patients at 6- and 12-month follow-ups, respectively. The demographic and clinical characteristics of these participants at baseline are listed in Table 1. There were no significant differences in age, gender distributions, or creatine levels among AH patients, HDCs, and HCs. Compared to HDCs and HCs, AH patients had elevated levels of total bilirubin, ALT, and AST. HDCs and HCs had comparable levels of total bilirubin and AST, but HDCs had higher levels of ALT than HCs. AH patients had longer prothrombin time and higher Model for End-Stage Liver Disease (MELD) score than HDCs. AH patients and HDCs reported similar drinking patterns (total drinking days and average number of drinks per day) over the preceding 30 days. Characteristics of the AH patients who achieved complete alcohol abstinence for 6-month or 12-month intervals as compared to HCs are shown in Supplementary Table 1. There were no differences in age, gender distribution, and creatinine levels between HCs and AH patients at baseline or follow-ups. Compared to HCs, AH patients at baseline and 6-month follow-up had increased levels of total bilirubin, ALT, and AST. At 12-month follow-up, most of those liver biochemistries in the AH subjects were normalized, except for ALT level, which still exhibited a trending increase (*p* = 0.065). For the AH subjects, both the prothrombin time and MELD score showed significant improvement at either the 6- or 12-follow-up in comparison to the baseline values.

### AH patients had impaired biosynthetic switch from PLMs to SPMs

To elucidate the distinctions in the profiles of circulating PLMs and SPMs between AH patients and HDCs as well as HCs, we first performed LC-MS/MS to quantify plasma levels of LMs and their precursors. LC-MS/MS offers numerous analytical advantages, including an extended linear dynamic range, the capability to simultaneously quantify multiple metabolic analytes, high accuracy and precision due to the use of internal standards, and the avoidance of the necessity for immunological reagents^52^. Our LC-MS/MS analysis included 13 pro- and anti-inflammatory LMs and 3 SPM intermediates in the ω-6 and ω-3 PUFA pathways, including 3 LMs (LTB4, LXA4, and LXB4) in the ω-6 pathway, RvE1 and its precursor 18-HEPE in the EPA pathway, and 11 SPMs (RvD1-5, PD1, PDX and their precursor 17-HDHA; MaR1, MaR2 and the MaR precursor 14-HDHA) in plasma samples from 10 AH patients, 10 HDCs, and 10 HCs. As shown in Supplementary Table 2, only 6 LMs were consistently detected in all 30 plasma samples, including LTB4, the RvE precursor 18-HEPE, RvD1/RvD2 and their precursor 17-HDHA, and MaR precursor 14-HDHA. The remaining 10 LMs were detected in only a small subset of the 30 samples, ranging from 0-10. Those 10 LMs were not further analyzed. We also quantified levels of 3 PLMs (LTB4, PGD2, and PGE2) and 4 SPMs (LXA4, RvE1, RvD2, and MaR1) in a larger set of plasma samples using ELISAs. We grouped the comparisons of the LM levels among the AH patients, HDCs, and HCs according to the AA, EPA, and DHA metabolic pathways.

In the ω-6 AA pathway (simplified schematic representation shown in Fig. 1A), the LC-MS/MS results revealed that AH patients had markedly higher plasma levels of the proinflammatory LTB4 (Fig. 1B), which was corroborated by the ELISA results (Fig. 1C). LC-MS/MS, but not ELISA, detected a higher LTB4 level in HDCs when compared with HCs (Fig. 1B, 1C). The PLM PGD2 levels were notably elevated in AH patients when compared to HCs and HDCs, whereas HDCs had similar levels of PGD2 to HCs (Fig. 1D). The levels of the PLM PGE2 were not different among AH patients, HDCs, and HCs (data not shown). Two SPMs (LXA4 and LXB4) in the AA pathway were detected in only a subset of plasma samples by LC-MS/MS (2 for LXA4 and 8 for LXB4) (Table S2). However, ELISA consistently detected LXA4 in all plasma samples tested, revealing that LXA4 levels were significantly reduced in AH patients compared to either HCs or HDCs (Fig. 1E). As the proinflammatory LTB4 and the anti-inflammatory LXA4 are derived from a common precursor LTA4, we also compared the LTB4/LXA4 ratio among AH patients, HDCs, and HCs. In line with a higher level of LTB4 and a reduced level of LXA4 in AH patients, the LTB4/LXA4 ratio was highly elevated in AH patients relative to HDCs and HCs (Fig. 1F). No difference in LXA4 or LTB4/LXA4 ratio was found between HDCs and HCs (Fig 1E, 1F).

**Figure 1.**
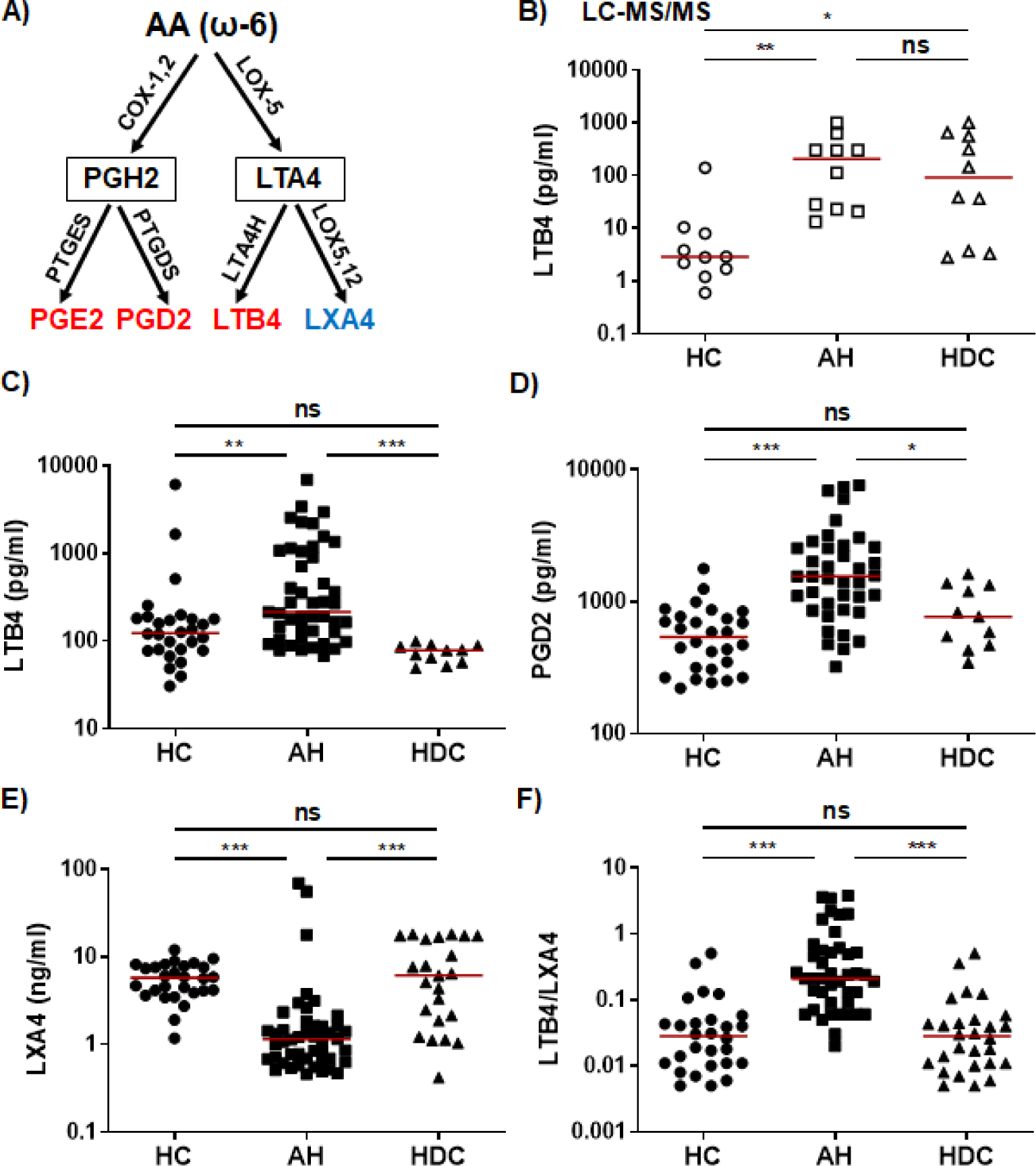
Dysregulated lipid mediators from the ω-6 AA pathway in AH patients. **A)** Simplified schematic representation of the ω-6 AA metabolic pathway. Key biosynthetic enzymes are shown next to arrows, lipid intermediators in box, and proinflammatory lipid mediators in red and anti-inflammatory lipid in blue. **B-F)** Scatter plots showing plasma levels of LTB4 (**B, C**), PGD2 (**D**), LXA4 (**E**), and LTB4/LXA4 ratio in HCs, AH patients, and HDCs. Concentrations were measured by LC-MS/MS (**B**; open symbols) or ELISA (**C-E**; filled symbols). Kruskal-Wallis test with Dunn’s correction for pairwise comparison among AH, HDC, and HC. AA, arachidonic acid; COX, cyclooxygenase; LOX, lipoxygenase; PTGDS, PGD2 synthetase; PTGES, PGE2 synthetase; AH, alcoholic hepatitis; HDC, heavy drinking control; HC, healthy control; \**p* < 0.05; \*\**p* < 0.01; \*\*\**p* < 0.001; ns, not significant.

The ω-3 SPMs, including Rvs, protectins, and MaRs, are derived from ω-3 PUFAs such as EPA and DHA through a series of enzymatic reactions involving 5-/12-/15-LOX. In the ω-3 EPA pathway (simplified schematic representation shown in Fig. 2A), plasma levels of the RvE intermediate 18-HEPE were significantly lower in AH patients compared to HCs as detected by LC-MS/MS (Fig. 2B). RvE1 was barely detectable by LC-MS/MS (Table S2). However, ELISA was able to detect RvE1 in all plasma samples, showing that AH patients had significantly higher levels than HCs or HDCs (Fig. 2C). In the ω-3 DHA pathway (simplified schematic representation shown in Fig. 3A), LC-MS/MS was able to detect the precursors to RvD, protectins, and MaRs (17-HDHA and 14-HDHA, respectively). Both precursors were significantly decreased in AH patients compared to HCs (Fig. 3B, 3C). In addition, HDCs also had lower levels of 17-HDHA levels than HCs (Fig. 3B). LC-MS/MS was able to detect RvD1 and RvD2 in all samples (Table S2). There were no significant differences in the levels of RvD1 and RvD2 among AH patients, HDCs, and HCs (Fig. 3D and data not shown). Of note, RvD1 levels in AH patients displayed a trend towards lower values compared to HCs (*p* = 0.066). MaR1 and MaR2 were not detected in most samples by LC-MS/MS. MaR1 was detectable by ELISA, but no discernible changes were observed in AH patients compared to HDCs and HCs (data not shown). Taken together, the results from our LC-MS/MS and ELISA analyses collectively suggest a pronounced imbalance in LM production in AH patients. This imbalance appears to skew towards elevated levels of PLMs and reduced levels of anti-inflammatory SPMs and their precursors when compared to HDCs and HCs.

**Figure 2.**
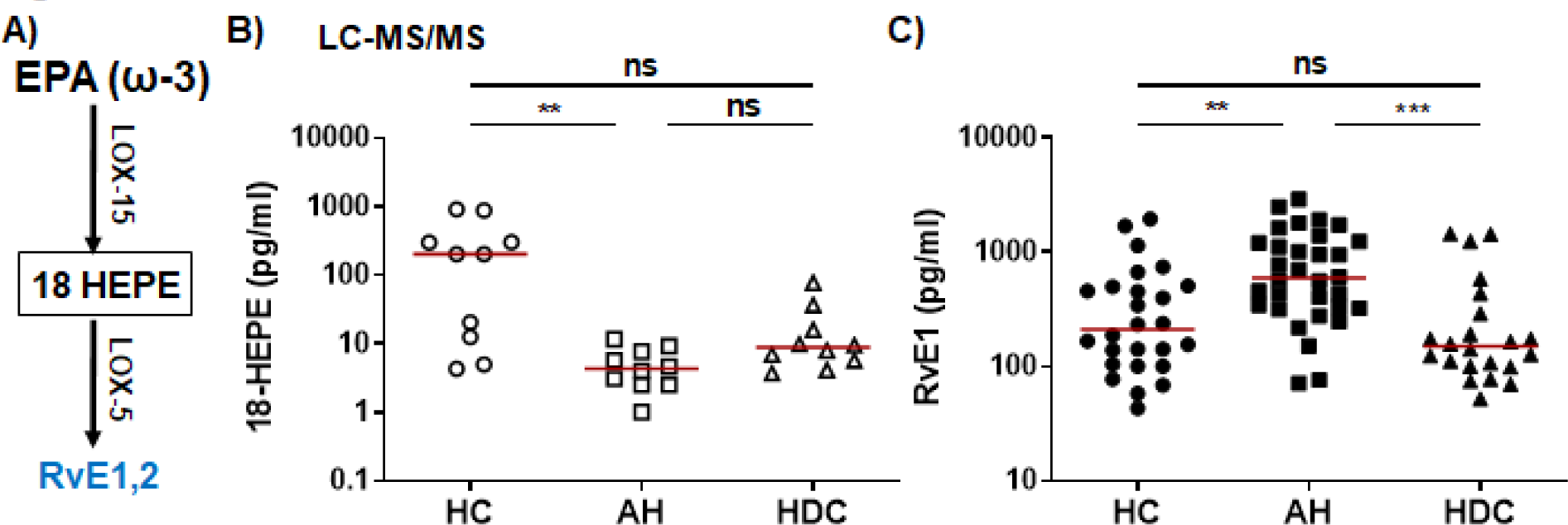
Dysregulated RvE1 and its precursor 18-HEPE from the ω-3 EPA pathway in AH patients. **A)** Simplified schematic representation of the ω-3 EPA metabolic pathway. Key biosynthetic enzymes are shown next to arrows, lipid intermediator in box, and anti-inflammatory SPMs (RvE series) in blue. **B, C)** Scatter plots showing plasma levels of 18-HEPE (**B**) and RvE1 (**C**) in HCs, AH patients, and HDCs. Concentrations were measured by LC-MS/MS (**B**; open symbols) or ELISA (**C**; filled symbols). Kruskal-Wallis test with Dunn’s correction for pairwise comparison among AH, HDC, and HC. EPA, eicosatetraenoic acid; LOX, lipoxygenase; AH, alcoholic hepatitis; HDC, heavy drinking control; HC, healthy control; \*\**p* < 0.01; \*\*\**p* < 0.001; ns, not significant.

**Figure 3.**
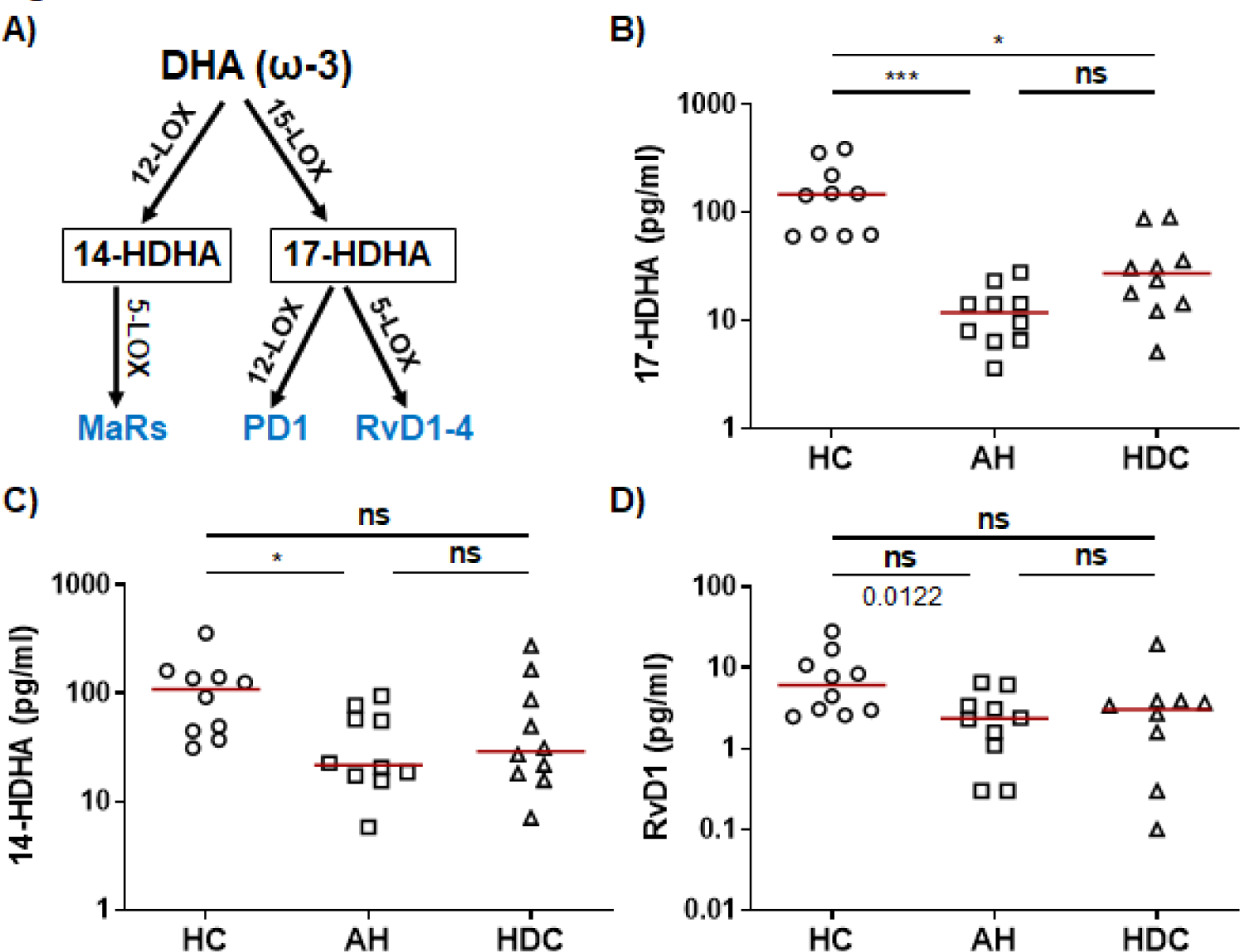
Dysregulated precursors of RvDs/PDs and MaRs from the ω-3 DHA pathway in AH patients. **A)** Simplified schematic representation of the ω-3 DHA metabolic pathway. Key biosynthetic enzymes are shown next to arrows, lipid intermediator in box, and anti-inflammatory SPMs in blue. **B-D)** Scatter plots showing LS-MS/MS quantification of plasma levels of RvD/PD precursor 17-HDHA (**B**), MaR precursor 14-HDHA (**C**), and RvD1 (**D**) in HC, AH patients, and HDCs). Kruskal-Wallis test with Dunn’s correction for pairwise comparison among AH, HDC, and HC. DHA, docosahexaenoic acid; LOX, lipoxygenase; AH, alcoholic hepatitis; HDC, heavy drinking control; HC, healthy control; \**p* < 0.05; \*\*\**p* < 0.001; ns, not significant.

### Alcohol abstinence reversed defective production of LMs

Next, we conducted a longitudinal assessment to determine whether the 3 highly dysregulated LMs (PGD2, LTB4, and LXA4) were changed in AH patients who achieved complete abstinence at follow-ups, using ELISA assays. Compared to the baseline values, PGD2 and LTB4 levels were significantly reduced (Fig. 4A-D), while changes in LXA4 levels remained relatively stable and did not show significant changes at either the 6- or 12-month follow-up (Fig. 4E, 4F). In addition, we determined whether the levels of those LMs were normalized at the follow-ups. At follow-ups, elevated levels of PGD2 and LTB4 at baseline had regressed to levels comparable to those detected in HCs, while the reduced levels of LXA4 had increased to levels analogous to those of HCs (Fig. S1). Thus, the levels of PGD2, LTB4, and LXA4 are normalized in abstinent AH patients over the course of 6- and 12-month follow-ups.

**Figure 4.**
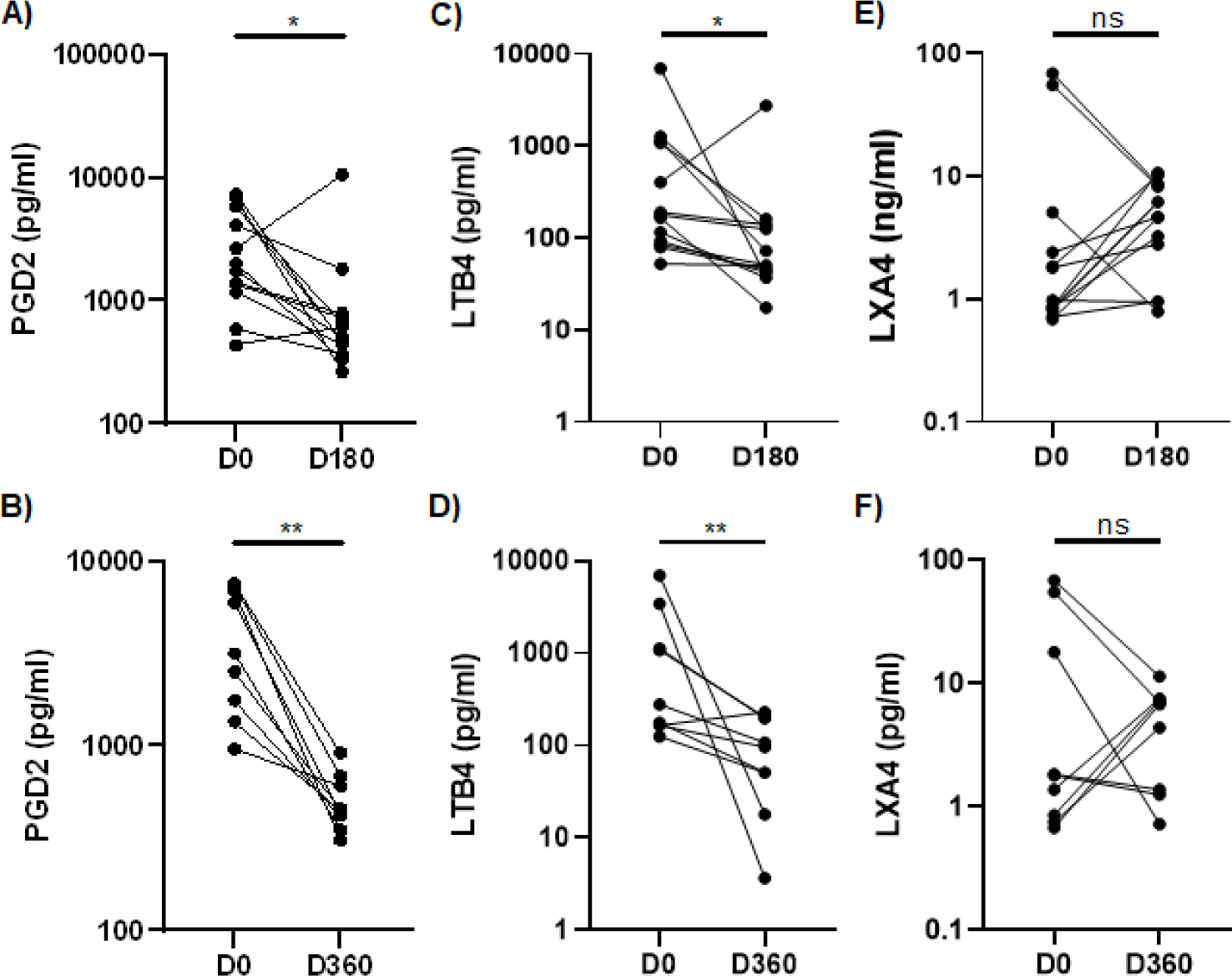
Longitudinal changes in plasma levels of PGD2, LTB4, and LXA4 in AH patients at 6-month or 12-month follow-ups. Scatter plots comparing levels of PGD2 (**A, B**), LTB4 (**C, D**), and LXA4 (**E, F**) at 6-month (D180) or 12-month (D360) follow-ups with baseline values (D0) in AH patients who were abstinent during the follow up intervals. Wilcoxon matched-pairs signed rank test was used to compare the differences between D0 and D180 (n=13) or D360 (n=9). \**p* < 0.05; \*\**p* < 0.01; ns, not significant. AH, alcoholic hepatitis.

### Data mining for gene expression of critical enzymes involved in LM production in AH patients

To unravel the underlying mechanisms contributing to the dysregulated production of LMs in AH patients, we conducted a comprehensive analysis of three publicly available RNA-seq databases (GSE143318, GSE142530, GSE155907). Our objective was to assess the differential expression of genes that encode several critical enzymes involved in LM production within liver tissue from AH patients and HCs serving as liver donors. The combined dataset encompassed RNA sequences from 20 AH patients and 21 HCs. We observed that the expression of COX-1 or COX-2, which converts the ω-6 AA PUFA to the precursor of PGE2 and PGD2 (PGH2), was not different between AH patients and HCs (data not shown). However, expression of the downstream PGD2 synthetase (PTGDS) and PGE2 synthetase (PTGES) were significantly upregulated in the liver tissue from AH patients (Fig. S2 and data not shown). In addition, the gene encoding the enzyme LTA4 dehydrogenase (LTA4H), responsible for converting LTA4 into LTB4 was also elevated in AH patients (Fig. S2B). The genes encoding for 5-LOX, an enzyme involved in the production of both PLMs (LTB4) and SPMs in the AA (LXA4 and LXB4) and the ω-3 pathways, were highly upregulated in the liver tissue from the AH patients (Fig. S2C). The gene encoding for 12-LOX that is essential for generating SPMs from both ω-6 and ω-3 pathways were also significantly enhanced in the liver tissue of AH patients (Fig. S2D). On the other hand, expression of 15-LOX, encoding for another critical LOX involved in SPM production, exhibited a trend towards higher in AH patients compared to HCs (*p* = 0.07 and data not shown). Together, liver tissue from AH patients exhibits a distinct gene expression profile indicative of heightened production of LMs, especially PLMs.

Monocytes/macrophages and neutrophils play central roles in inflammation initiation and resolution by producing a plethora of proinflammatory and anti-inflammatory mediators, as well as phagocytosing cellular debris and injured cells^53–55^. We analyzed RNA expression of genes involved in LM production in peripheral blood monocytes (GSE135285) and neutrophils from AH patients (GSE171809). CD14^+^ monocytes from patients with severe AH (n=4) and HCs (n=6) had comparable expression levels of COX-1, COX-2, 5-LOX, 12-LOX, 15-LOX, PTGES, and LTA4H (data not shown), with exception of PTGDS, which showed a higher expression trend in the monocytes from AH patients (p=0.055). A recent study identified a unique and functionally exhausted subpopulation of neutrophils, referred to as low-density neutrophils (LDN), in the AH subjects that are different from the conventional high-density neutrophils (HDN)^56^. Interestingly, LDN from AH patients expressed notably higher levels of COX-1 and COX-2 than HDN in AH subjects and HCs. In contrast, the expression of these genes in HDN form AH subjects and HCs were comparable (Fig. S3A, S3B). Both 5-LOX and 12-LOX in LDN were downregulated in AH subjects compared to that from HCs. In addition, 12-LOX expression in HDN from AH subjects was also lower than those from HCs. However, 5-LOX levels in HDN were higher than both LDN and neutrophils from HCs. These data suggest that peripheral LDNs in AH patients possess the potential to produce higher levels of PLMs while generating lower levels of SPMs.

### Correlation of plasma levels of LMs with disease severity and inflammatory mediators in AH patients

To examine whether LMs might be linked to the pathogenesis of AH, we performed Spearman correlation analyses to assess their associations with AH disease severity markers, including creatinine, liver-related biochemical measurements (total bilirubin, AST, and ALT), prothrombin time, and MELD score in AH patients. Plasma levels of PGD2, PDE2, LTB4, LXA4, MaR1, and RvE1 as determined by ELISA were used for these analyses. The results of these correlations are summarized in Table 2. Upregulated PGD2 positively correlated with total bilirubin and displayed a trend towards correlation with creatinine and AST (*p* = 0.07 for both). MaR1 exhibited a negative correlation with ALT. LTB4 and LXA4, as well as PGE2 and RvE1, did not correlate with any of these markers of disease severity (Table 2 and data not shown). However, the elevated LTB4/LXA4 ratio displayed a positive correlation with the MELD score (Table 2), suggesting this specific ration may hold potential as an indicator of disease severity in AH patients.

**Table 2.**
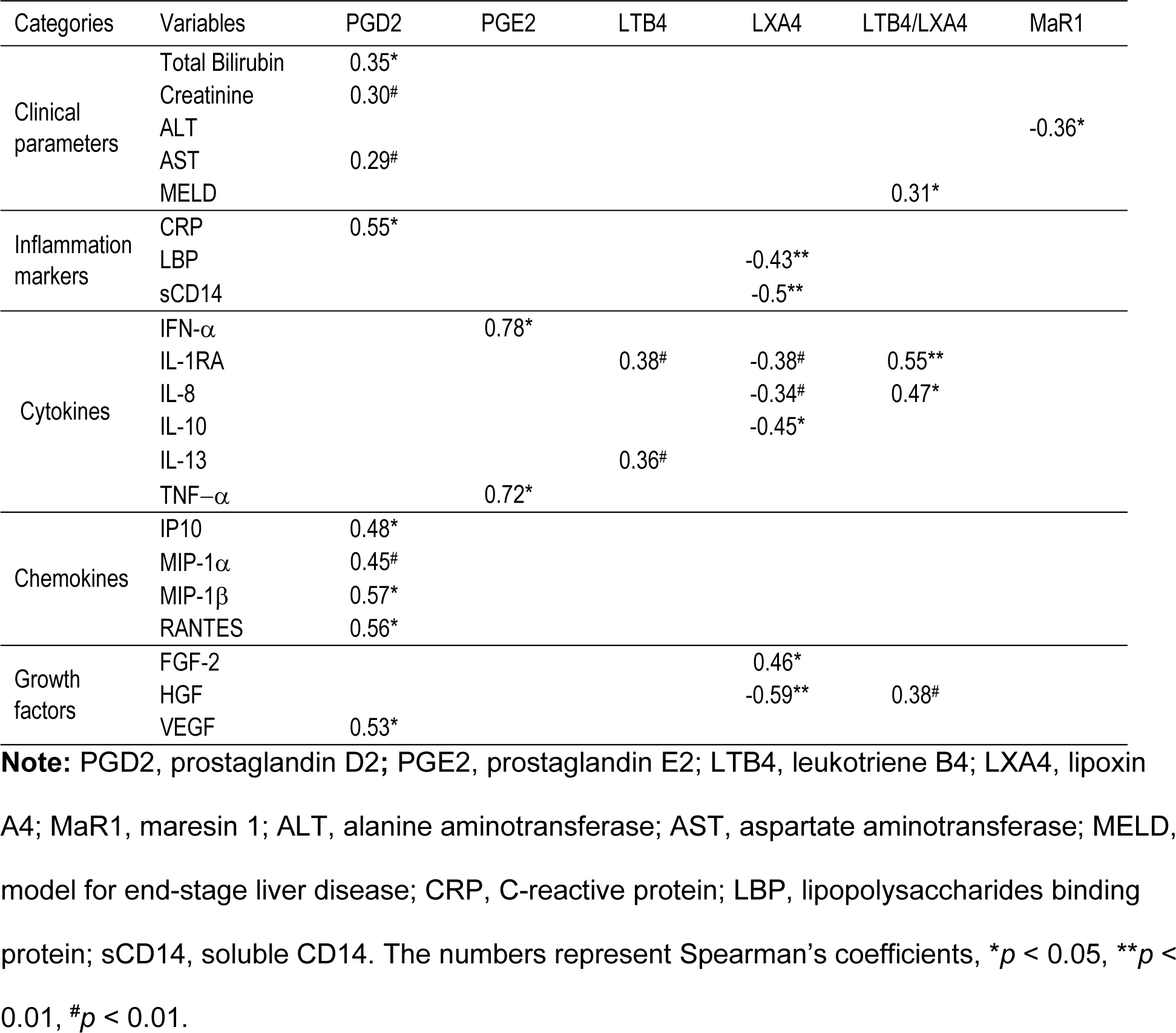
Correlations of altered lipid mediators with clinical parameters and inflammatory mediators in AH patients.

| Categories | Variables | PGD2 | PGE2 | LTB4 | LXA4 | LTB4/LXA4 | MaR1 |
| --- | --- | --- | --- | --- | --- | --- | --- |
| Clinical parameters | Total Bilirubin | 0.35* |  |  |  |  |  |
|  | Creatinine | 0.30<sup>#</sup> |  |  |  |  |  |
|  | ALT |  |  |  |  |  | -0.36* |
|  | AST | 0.29<sup>#</sup> |  |  |  |  |  |
|  | MELD |  |  |  |  | 0.31* |  |
| Inflammation markers | CRP | 0.55* |  |  |  |  |  |
|  | LBP |  |  |  | -0.43** |  |  |
|  | sCD14 |  |  |  | -0.5** |  |  |
| Cytokines | IFN-α | | 0.78* | | | | |
|  | IL-1RA |  |  | 0.38<sup>#</sup> | -0.38<sup>#</sup> | 0.55** |  |
|  | IL-8 |  |  |  | -0.34<sup>#</sup> | 0.47* |  |
|  | IL-10 |  |  |  | -0.45* |  |  |
|  | IL-13 |  |  | 0.36<sup>#</sup> |  |  |  |
| | TNF-α | | 0.72* | | | | |
| Chemokines | IP10 | 0.48* |  |  |  |  |  |
| | MIP-1α | 0.45<sup>#</sup> | | | | | |
| | MIP-1β | 0.57* | | | | | |
|  | RANTES | 0.56* |  |  |  |  |  |
| Growth factors | FGF-2 |  |  |  | 0.46* |  |  |
|  | HGF |  |  |  | -0.59** | 0.38<sup>#</sup> |  |
|  | VEGF | 0.53* |  |  |  |  |  |

**Note:** PGD2, prostaglandin D2; PGE2, prostaglandin E2; LTB4, leukotriene B4; LXA4, lipoxin A4; MaR1, maresin 1; ALT, alanine aminotransferase; AST, aspartate aminotransferase; MELD, model for end-stage liver disease; CRP, C-reactive protein; LBP, lipopolysaccharides binding protein; sCD14, soluble CD14. The numbers represent Spearman's coefficients, \* $p < 0.05$ , \*\* $p < 0.01$ , <sup>#</sup> $p < 0.01$ .

AH patients are in a hyperinflammatory state, primarily driven by alcohol-induced MT^8,9,12,13^. Inflammation and MT play pivotal roles in the development and progression of AH and also regulate the biosynthesis of LMs^57–59^. We first analyzed the associations of the 5 LMs (PGD2, PDE2, LTB4, LXA4, MaR1, and RvE1) with upregulated markers of systemic inflammation (CRP) and MT (LPS, LBP, sCD14, and sCD163)^10,11^. The significant correlations were summarized in Table 2. PGD2 had a significant positive correlation with plasma levels of CRP, whereas LXA4 correlated negatively with LBP and sCD14. None of the 5 LMs correlated with circulatory levels of LPS or sCD163 (data not shown). Furthermore, we performed a correlation analysis between plasma levels of these LMs and inflammatory cytokines, chemokines, and growth factors as quantified through a 45plex immunoassay. Upregulated PGD2 levels positively correlated with 3 chemokines (IP10, MIP-1b, and RANTES) and the growth factor VEGF and had a clear trend to correlate with another chemokine MIP-1a (*p* = 0.051). Elevated TLB4 trended to positively correlate with the IL-1 receptor antagonist (IL-1RA) (*p* = 0.06) and the IL-13 cytokine (*p* = 0.078). PGE2 showed positive correlations with the proinflammatory cytokines IFN-a and TNF-a. Downregulated LXA4 negatively correlated with the anti-inflammatory cytokine IL-10 and the growth factor HGF (hepatocyte growth factor).. LXA4 also trended to negatively correlated with IL-1RA and IL-8, both of which exhibited positive correlations with the elevated LTB4/LXA4 ratio. Of note, the inflammatory factors linked with the dysregulated LMs in AH patients were all significantly elevated in AH patients compared with HC and/or HDC (Supplemental Table 3). These results suggest that dysregulated production of LMs is intricately linked with dysregulated production of inflammatory cytokines, chemokines, and growth factors, and potentially contributing to the pathogenesis of AH.

## Discussion

AH is a severe and progressive liver and systemic inflammatory disease. Our recent investigations, consisting of cross-sectional and longitudinal studies within a comprehensive multicenter project (TREAT, U01AA021840), have unveiled crucial insights into the inflammatory responses, microbial translocation, the activation of immune cells and endothelial cells (ECs), and intestinal epithelium damage^4,10,11,51^. Our results reveal that even after 12 months of abstinence from alcohol, AH patients continue to exhibit significantly elevated levels of inflammatory mediators, such as the proinflammatory cytokines IL-8, TNF-α, IL-18, and IL-23^4^. These results strongly imply that alcohol not only triggers inflammation but also impairs the resolution of inflammation. However, little research has been conducted to study the biological events that regulate the resolution of inflammation in ALD such as AH. In this study, we used LS-MS/MS and ELISA to profile various LMs in the peripheral blood, encompassing both PLMs and SPMs, in AH patients, HDCs, and HCs. Th primary objective was to understand the biological processes orchestrating the resolution of inflammation and the restoration of normal metabolism and tissue homeostasis in AH patients as compared to heavy drinkers without AH. We found that AH patients had higher circulating levels of two PLMs (PGD_2_ and LTB_4_) and the SPM RvE1 compared to HDCs and/or HCs (Figs. 1B-D, 2C). In contrast, the SPMs LXA4 (Fig. 1E) and RvD1 (Fig. 3D), along with the precursors to RvEs (18-HEPE), RvDs (17-HDHA), and MaRs (14-HDHA) (Fig. 2B, 3B, 3C) within the ω-3 pathway, were significantly reduced in AH subjects. Notably, the plasma LM profile in HDCs remained largely unaffected when compared to HC, except for higher levels of LTB4 (Fig 1B) and lower levels of 17-HDHA (Fig. 3B) detected using LC-MS/MS. Intriguingly, some PLMs and SPMs correlated AH disease severity, clinical parameters, and several inflammatory cytokines positively and negatively, respectively (Table 2). The dysregulation in LMs was reversed with alcohol abstinence at 6- and 12-month follow-ups (Fig. 4, Fig. S1), coincident with the normalization of clinical parameters (Supporting Table S1). Although HDCs had no overt clinical symptoms, they still exhibited some alterations in circulating levels of PLMs and SPMs (Table 1, Fig. 1B, 3B).

The failure to transition from the production of PLMs to SPMs leads to defective resolution of inflammation, contributing to the progression of various chronic inflammatory disorders^60,61^. In individuals with alcohol use disorder (AUD) or AH, serum profiles of bioactive lipid metabolites derived from the oxidation of ω-6 and ω-3 PUFAs are profoundly altered as determined by LC-MS/MS^62^. This study showed that serum level of LTB4, one of PLMs in ω-6 pathway, is significantly higher and positively correlated with MELD score^62^. Our study corroborates these findings, revealing significantly elevated plasma levels of LTB4 in AH patients (Fig. 1), but no no direct correlation between upregulated LTB4 level and MELD score was ibserved in our AH subjects. However, we did identify a positive correlation between the LTB4/LXA4 ratio and the MELD score (Fig. 1F). Additionally, the highly elevated PGD2 correlated with several aspects of AH disease severity, which might contribute to hepatic information in AH. LTB4 is enzymatically generated from AA through the action of 5-LOX and LTA4H, while PGD2 is biosynthesized from AA via COX-1 and COX-2, as well as terminal PGDS. In line with the heightened plasma levels of PGD2 and LTB4 in our AH patient cohort, an analysis of three publicly available RNA-seq databases of liver tissue from AH subjects revealed a significant upregulation in the gene expression of PGDS and LTA4H (Supporting Fig. S2A, S2B).

Circulatory LX levels exhibit a marked reduction in various inflammatory diseases^63–65^. Intriguingly, the reduced circulating levels of LXA4 in the AH subjects did not align with the higher gene expression of both *ALOX5* and *ALOX12* critical for LXA4 production in liver tissue from AH patients (Supporting Fig. S2C, S2D). This discrepancy could be due to multiple reasons, such as high hepatic infiltration of ALOX-expressing inflammatory leukocytes, translation efficiency, the differences in cell sources, subcellular location and inactivation of the enzymes, different regulators of ALOX enzyme activity, and tissue/cells outside liver contribute to LX production in circulation^66^. ALOX5 is the central enzyme for the biosynthesis of the PLM leukotrienes and the SPM LXs and Rvs. Its association with the regulatory protein (5-lipoxygenase-activating protein (FLAP) determines the preferential synthesis of either PLMs or SPMs. Our data mining revealed that LDN, a subpopulation of peripheral blood neutrophils, expressed a lower level of ALOX5 as well as ALOX12 than conventional HDN from HCs and AH subjects (Supporting Fig. S3C, S3D), suggesting that these unconventional neutrophil subpopulation might contribute to the reduced production of LXA4 in AH patients.

ω-3 SPMs, encompassing DHA-derived MaRs, protectins, and D-series Rvs, as well as EPA-derived E-series Rvs, play a pivotal role in regulating inflammation and restoring homeostasis. The levels of these ω-3 SPMs in circulation are frequently altered in chronic inflammatory diseases^67^. Our LS-MS/MS results revealed a significant decrease in the levels of precursors for Rvs, protectins, and MaRs of SPMS (18-HEPE, 17-HDHA, and 14-HDHA) in AH patients (Fig. 3B, 3C). Notably, these SPM intermediates, 17-HDHA and 18-HEPE, have been used as markers for the activation levels of the MaR-, protectin-, and Rv-producing pathways, respectively^87–89^. In addition, plasma level of RvD1 was lower in AH patients (Fig. 3D). RvD1 is known for its protective effects in various liver diseases, such as different forms of liver injury, liver fibrosis, and liver cancer^68^. The reduced level of RvD1 may contribute to hepatic and systemic inflammation in AH.

AH patients are in a highly inflammatory and immune-dysregulated state^4^, which underscores the intricate interplay between proinflammatory and anti-inflammatory LMs. Our results revealed that two PLMs (PGD2 and PEG2) positively correlated with the widely recognized inflammation marker CRP and several proinflammatory cytokines, chemokines, and growth factors, while the SPM LXA4 exhibited an inverse correlation with MT markers and upregulated IL-10 and HGF in AH patients (Table 2). Proinflammatory cytokines play a reciprocal role in upregulating the expression of enzymes responsible for LM biosynthesis^64^. For example, both IL-4 and IL-13 can induce 15-ALOX expression in human monocytes^69,70^. GM-CSF can elevate the expression of 5-ALOX in neutrophils^71^. The imbalanced production of PLMs and SPMs in AH likely plays a role in enhancing production of proinflammatory cytokines and other inflammatory mediators that can feedback to further alter LM profiles by modulating enzymes involved in LM biosynthesis.

Currently, there are no effective medical interventions available for AH. Alcohol abstinence remains a cornerstone of treatment for ALD, which significantly improves the disease outcomes, but typically does not lead to complete recovery for most ALD patients, consistent with our clinical observations(Supporting Table S1)^51^. Despite alcohol abstinence, our previous studies involving this cohort of AH patients revealed the persistence of numerous up-regulated inflammatory factors, including proinflammatory cytokines (IL-8, IL-18, IL-23, and TNF-α), EC activation markers (sCD146, sICAM, and sVCAM), soluble immune checkpoints (sCD27, sCD40, sHVEM, and sTIM3), and the intestinal epithelium damage markers (REG3a and TFF3)^4,10,51,72^. In our current study, we found that three highly dysregulated LMs (PGD2, LTB4, and LXA4) were mostly normalized in AH subjects who abstained from alcohol (Supporting Fig. S1). While it remains unknown whether other LMs, especially SPMs, experience a similar restoration, our data strongly suggest that LMs may exhibit a more pronounced responsiveness to the cessation of alcohol consumption.

In conclusion, our investigation unveils a substantial dysregulation of peripheral blood LMs, encompassing a broad spectrum of PLMs, SPMs, and their precursors in AH patients. These abnormalities were largely reversed in AH subjects who underwent alcohol abstinence. Moreover, our study revealed correlations between altered LM levels and key clinical indicators of disease severity, as well as inflammatory factors in AH patients. Currently, there is no effective medical treatment available for AH^90^. Corticosteroids remain the mainstay of treatment for severe AH^90,91^, but cause serious side effects and immunosuppression^90^. Thus, there is an urgent need to identify safer and more effective therapeutics. A promising approach involves harnessing the body’s natural resolution process of inflammation for therapeutic purposes. SPMs actively exert potent dual anti-inflammatory and pro-resolving effects without causing immunosuppression^42,45,59^. Furthermore, SPMs contribute to the restoration of tissue metabolism and homeostasis. Therefore, SPMs emerge as a valuable and innovative tool for preventing, ameliorating, and treating AH and other forms of ALD. In a mouse model of AH, 12-/15-LOX deficiency exacerbated hepatic and systematic inflammation and increased liver disease severity^73^. Remarkably, this pathological state could be effectively mitigated through the exogenous supplementation of LXA4^73^. Our study, demonstrating the skewed production of PLMs and SPMs in AH patients, adds further support to the development of inflammation resolution-based strategies for the prevention, amelioration, and treatment of AH.

## Data Availability

All data produced in the present study are available upon reasonable request to the authors

https://medicine.iu.edu/faculty/6507/yu-andy

## Abbreviations

AA: arachidonic acid
AH: alcoholic hepatitis
ALA: alpha linolenic acid
ALD: alcoholic liver diseases
ALT: alanine aminotransferase
AST: aspartate aminotransferase
COX: cyclooxygenase
DHA: docosahexaenoic acid
EPA: eicosatetraenoic acid
GI: gastrointestinal
HC: healthy control
HDC: heavy drinking control
HDHA: hydroxy docosahexaenoic acid
HEPE: hydroxyeicosapentaenoic acid
HGF: hepatocyte growth factor
INR: international normalized ratio
LA: linolenic acid
LBP: lipopolysaccharides binding protein
LC-MS/MS: Liquid chromatography and tandem mass spectrometry
LM: lipid mediator
LOX: lipoxygenase
LPS: lipopolysaccharides
LT: leukotriene
LTA4H: LTA4 dehydrogenase
LX: lipoxin
MAIT cells: mucosal-associated invariant T cells
MaR: maresin
MELD: model for end stage liver disease
M/M*Φ*: monocytes/macrophages
MT: microbial translocation
NIAAA: National Institute on Alcohol Abuse and Alcoholism
OS: oxidative stress
PD: protectin
PG: prostaglandin
PLM: pro-inflammatory lipid mediator
PTGDS: PGD2 synthetase
PTGES: PGE2 synthetase
PUFA: poly-unsaturated fatty acid
ROS: reactive oxygen species
Rv: resolving
sCD14: soluble CD14
sCD163: soluble CD163
SPM: specialized pro-resolving mediator
TREAT: Translational Research and Evolving Alcoholic Hepatitis Treatment
Tx: thromboxane

## Acknowledgments

We thank the members of the TREAT consortium (Translational Research and Evolving Alcoholic Hepatitis Treatment) at Indiana University (NIAAA AA021883), Mayo Clinic (NIAAA AA021788), and Virginia Commonwealth University (NIAAA AA021891) Special appreciation extends to our collaborators at NIAAA, notably Dr. Svetlana Raedeva, for their invaluable contributions. We also acknowledge the unwavering support of the National Institute on Alcohol Abuse and Alcoholism (NIAAA U01 AA021840). This work was also funded by NIAAA grant (UH2/UH3AA026218 to QY) and the grant (OPP1035237 to QY) from the Bill & Melinda Gates Foundation.

## Specific author contributions

WL: conceptualized the study, designed and performed the experiments, performed data analysis, interpreted data, and wrote the manuscript. YX: designed and performed the experiments, performed data analysis. JY and FS: performed the experiments. AJS, VHS, and NC: provided clinical samples and feedback for the project, and critically reviewed the manuscript. QY: conceptualized the study, obtained funding for the study, critically reviewed, and finalized the manuscript.

