## supplementary figures and supplementary Tables for "Disrupted balance between pro-inflammatory lipid mediators and anti-inflammatory specialized pro-resolving mediators is linked to hyperinflammation in patients with alcoholic hepatitis": SPM2023SupFig.pdf

### Figure S1

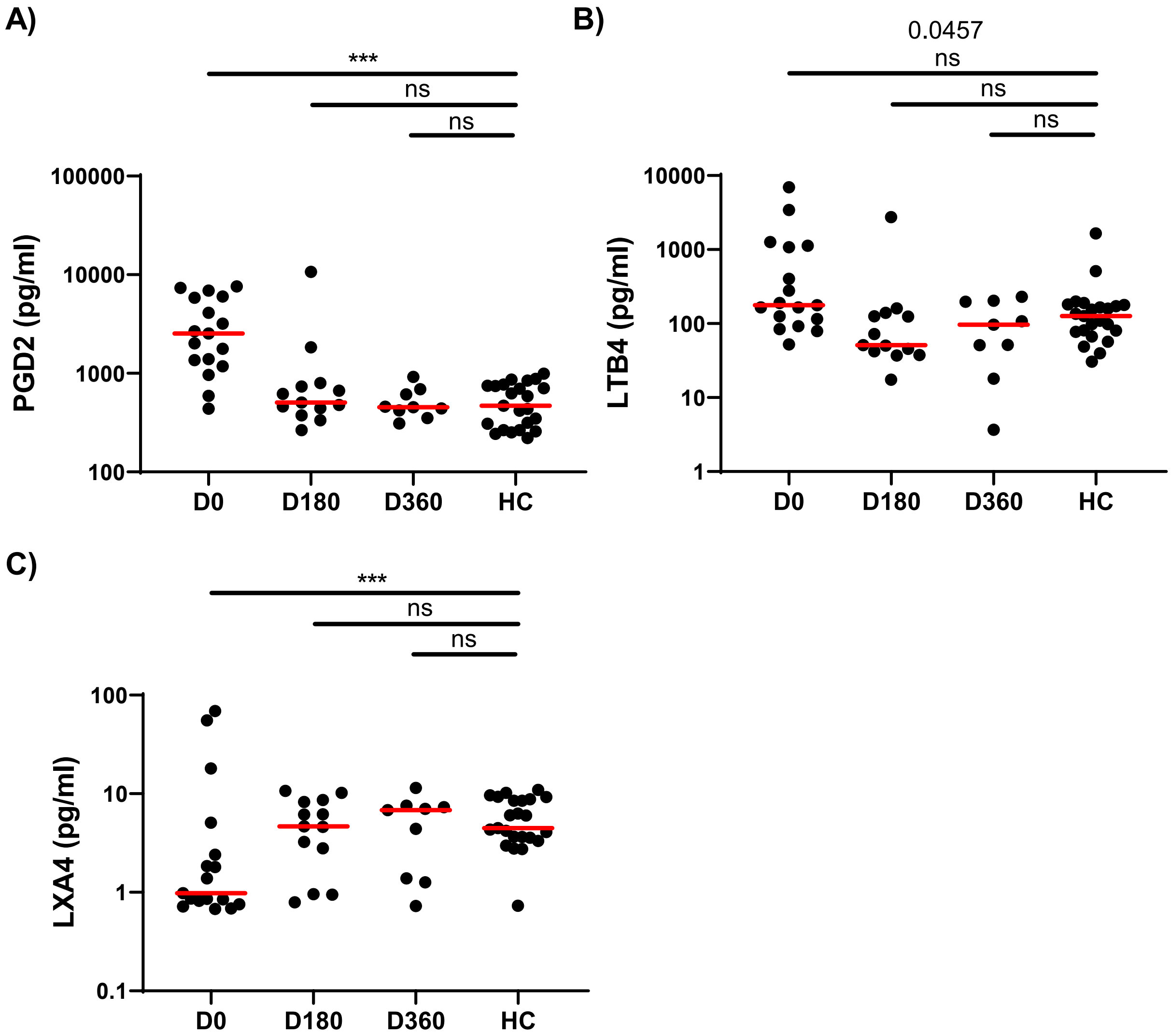


**Figure S1. Dysregulated pro- and anti-inflammatory lipid mediators were normalized in 6- and 12-month follow-up samples from AH patients who abstinent from alcohol.** Scatter plots showing plasma levels of PGD2 (**A**), LTB4 (**B**), and LXA4 (**C**) in AH patients at enrollment (D0), 6-month follow-up (D180), 12-month follow-up (D360), and healthy controls (HCs). Kruskal-Wallis test with Dunn's correction for comparisons of D0, D180, and D360 with HCs. ***$p <$ 0.001; ns, not significant.

### Figure S2

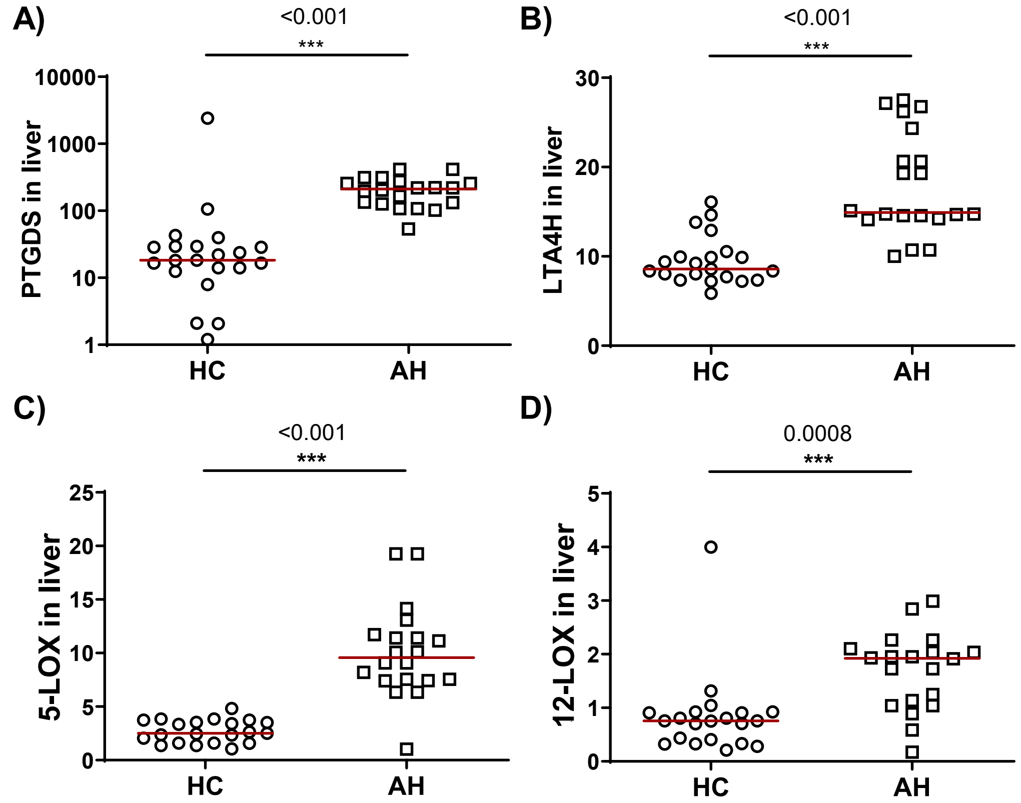


**Figure S2. Differential expression of genes involved in production of lipid mediators in liver tissue from AH patients and HCs.** RNA expression levels were pooled from liver tissue RNA-seq databases GSE143318, GSE152530, and GSE155907. Data are presented as TPM normalized expression values. TPM, transcripts per kilobase million; HC, healthy control; AH, patient with alcoholic hepatitis. Two-tailed t test was used to calculate differences between AH patients and HCs. ***$p < 0.001$.

Figure S3

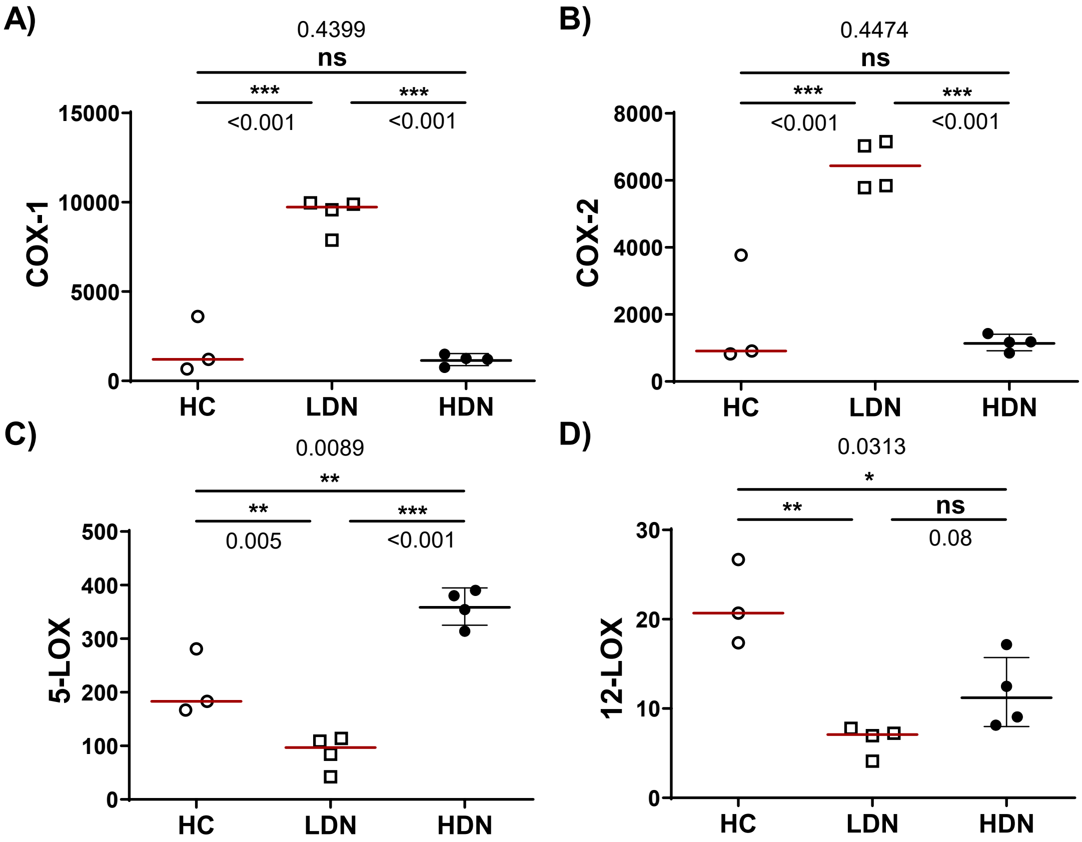


**Figure S3. Differential expression of genes involved in production of lipid mediators in peripheral blood neutrophils in AH patients and HCs.** RNA expression levels were extracted from neutrophil RNA-seq database GSE1710809. Data are presented as TPM normalized expression values. TPM, transcripts per kilobase million; HC, healthy controls; LDN, low density neutrophils; HDN, high density neutrophils; AH, alcoholic hepatitis. Ordinary one-way ANOVA with Holm-Sidak's multiple comparisons test was used for comparisons among HC, LDN, and HDN. $*p < 0.05$; $**p < 0.01$; $***p < 0.001$; ns, not significant.
