## supplementary figures and supplementary Tables for "Disrupted balance between pro-inflammatory lipid mediators and anti-inflammatory specialized pro-resolving mediators is linked to hyperinflammation in patients with alcoholic hepatitis": SPM2023SupplementalTables.pdf

**Supplemental Table 1. Comparison of the clinical characteristics of the longitudinal cohort of patients**

| Variables | AH at baseline (n=17) | AH at 6-month (n=13) | AH at 12-month (n=9) | HC (n=23) |
|---|---|---|---|---|
| Age (years) | 45 (33-53) | 40 (32-50) | 45 (37-54) | 42 (30-56) |
| Gender (% Male) | 53 | 62 | 44 | 60 |
| Creatinine (mg/dL) | 0.80 (0.69-1.34) | 0.70 (0.57-1.00) | 0.86 (0.60-1.14) | 0.86 (0.73-1.02) |
| Total bilirubin (mg/dL) | 14.3 (7.8-21.9)*** | 1.2 (0.7-4.6)** | 0.7 (0.5-1.4) | 0.4 (0.2-0.5) |
| ALT (IU/L) | 50 (26-65)*** | 26 (21-41)** | 24 (18-33) | 12 (8-18) |
| AST (IU/L) | 115 (82-147)*** | 34 (30-72)** | 28 (24-37) | 17 (13-22) |
| Prothrombin time (INR) | 1.84 (1.62-2.28) | 1.33 (1.09-1.54)## | 1.10 (1.03-1.43)## | |
| MELD score | 25 (20-30) | 10 (8-16)## | 9 (7-13)### | |

**Note:** Data are represented as median and interquartile ranges and %. AH, patients with alcoholic hepatitis; HC, healthy controls; ALT, alanine aminotransferase; AST, aspartate aminotransferase; INR, international normalized ratio; MELD, model for end-stage liver disease. Kruskal-Wallis test with Dunn's correction was used for comparisons between HC and AH patients at baseline, 6-month, or 12-month follow-up. $^{**}p < 0.01$, $^{***}p < 0.001$. Kruskal-Wallis test with Dunn's correction for comparisons between AH patients at baseline and at 6-month or 12-month follow-up. $^{\#\#}p < 0.01$, $^{\#\#\#}p < 0.001$. Chi-square test for analysis of gender difference.

**Supplemental Table 2. Numbers of plasma samples with detectable levels of lipid mediators/precursors as measured by LS-MS/MS**

| | | HC (n=10) | AH (n=10) | HDC (n=10) |
|---|---|---|---|---|
| AA pathway | Leukotriene B4 | 10 | 10 | 10 |
| | Lipoxin A4 | 1 | 1 | 0 |
| | Lipoxin B4 | 5 | 1 | 2 |
| EPA pathway | 18-HEPE | 10 | 10 | 10 |
| | Resolvin E1 | 1 | 2 | 2 |
| DHA pathway | 17-HDHA | 10 | 10 | 10 |
| | Resolvin D1 | 10 | 10 | 10 |
| | Resolvin D2 | 10 | 10 | 10 |
| | Resolvin D3 | 3 | 1 | 0 |
| | Resolvin D4 | 3 | 4 | 3 |
| | Resolvin D5 | 2 | 4 | 1 |
| | Protectin 1 | 1 | 1 | 2 |
| | Protectin DX | 2 | 2 | 1 |
| | 14-HDHA | 10 | 10 | 10 |
| | Maresin 1 | 3 | 1 | 2 |
| | Maresin 2 | 0 | 0 | 0 |

**Note:** HC, healthy controls; AH, patients with alcoholic hepatitis; HDC, heavy drinking controls; AA, arachidonic acid; EPA, eicosatetraenoic acid; HEPE, hydroxyeicosapentaenoic acid; DHA, docosahexaenoic acid; HDHA, hydroxy docosahexaenoic acid.
